# Paradigms about the COVID-19 pandemic: knowledge, attitudes and practices from medical students

**DOI:** 10.1101/2020.05.21.20105858

**Authors:** Lincango-Naranjo Eddy, Solis-Pazmino Paola, Rodriguez-Villafuerte Santiago, Lincango-Naranjo Jose, Vinueza-Moreano Paul, Barberis-Barcia Giuseppe, Ruiz-Sosa Carlos, Rojas-Velasco Giovanni, Gravholt Derek, Espinoza-Suarez Nataly, Golembiewski Elizabeth, Soto-Becerra Percy

## Abstract

**Background:** As the disease caused by the new coronavirus has spread globally, economic instability in healthcare systems has been significant, and the lack of knowledge, positive attitudes and appropriate practices among people has led to the magnification of this disease. This reality is especially accentuated in Ecuador where, although many healthcare workers have been called to help in the regions most affected, the shortage of them combined with cultural and macroeconomic factors have led Ecuador to face the most aggressive outbreak in Latin America. In this context, the participation on the front line of final year medical students is indispensable. For that reason, appropriate training on COVID-19 for final year medical students is an urgent need that universities and health systems must guarantee. We aimed to describe the knowledge, attitudes and practices in Ecuadorian final year medical students in order to identify the knowledge gaps, perceptions and behavior patterns which could guide the design of better medical education curricula regarding COVID-19.

**Methods:** This was a cross-sectional 33-item online survey conducted between April 6 to April 20 assessing the knowledge, attitudes, and practices toward the diagnosis, treatment, prevention, and prognosis toward COVID-19 in Ecuadorian final year medical students. It was sent by email and by Facebook and WhatsApp.

**Results:** A total of 309 students responded to the survey. 88% of students scored high (≥ 70% correct) for knowledge of the disease. The majority of students were pessimistic about possible government actions, which is reflected in the negative attitude towards the control of COVID-19 in Ecuador and volunteering during the outbreak (77%, and 58% of the students, respectively). Moreover, 91% of students said they did not have adequate protective equipment or training in their health facilities.

**Conclusions:** The high level of students’ knowledge, with coexisting negative attitudes, suggests Ecuador has a capable upcoming workforce that could benefit from an opportunity to strengthen, improve and advance their training in preparation for COVID-19. Creating a national curriculum may be one of the most effective ways for all students to be trained, while simultaneously focusing on the students’ most pressing concerns. Hopefully with this, negative attitudes will improve and students will be better qualified.

## Introduction

The severe acute respiratory syndrome coronavirus 2 (SARS-CoV-2) is a respiratory virus that^1^ causes coronavirus disease 2019 (COVID-19), a potentially lethal infectious condition. Due to its high infectivity and transmissibility, SARS-CoV-2 has quickly caused a global pandemic ^2^ that started in China on November 2019. On February 29, 2020, the first known case of COVID-19 was reported in Ecuador^3^, and after only a few weeks Ecuador was facing the most aggressive COVID-19 outbreak in Latin America ^4,5^. The number of COVID-19 cases and deaths in Ecuador has gradually increased since February, with 29 559 cases out of 17 468 429 inhabitants and 2 127 deaths reported up to May 5 ^6,7^ making the incidence rate of cases per population in Ecuador one of the greatest in South America ^3,4^. This pandemic generated a worldwide lockdown, and several actions have been taken by national governments. In Ecuador, since March 15 the government has implemented shelter-in-place measures, closed schools, and shut down venues for social gathering, including restaurants, bars, movie theaters, concerts, sporting events, and others ^3^. However, these interventions have not been fully successful in stopping the spread of COVID-19, and the number of cases has continued to increase, with the Province of Guayas most affected. For that reason, many healthcare workers (HCWs) have been called to help the overwhelmed COVID-19 patients in this province.

HCWs are on the front line of COVID-19 outbreak and, as such, are at higher risk of contracting the virus. Around the world, a large number of them have been infected with COVID-19 ^8,9^ and thousands have died ^10^. In Ecuador alone, 1600 HCWs have been tested positive for the virus and 24 have died as of this writing ^6^. Being a HCW not only involve licensed professionals, such as doctors, nurses, pharmacists, and dentists but also medical students, specifically those who are in their last year doing a clinical internship. During this last stage of their medical training, Ecuadorian medical students participate in a wide range of key activities within the health system, such as the comprehensive care of people in their life, family and community cycle; the education aimed at health promotion and disease prevention; the administrative support in health units; and even in planning of health services, policies, programs and projects aimed at specific groups and community benefits. Thus, the last year is essential for students to integrate and consolidate the knowledge they have acquired during the previous years. They have little experience managing patients and can easily make mistakes. This lack of experience increases their chance of infection. If they are infected, the likelihood of developing deadly forms of COVID-19 is low, but it is high for developing an asymptomatic disease, and becoming vectors which exacerbate the epidemiological burden of the disease.

At present, since no treatment or vaccine is available for COVID-19 and patients are managed with supportive care, applying preventive measures to control COVID-19 is the most critical intervention. Organizations such as the CDC and WHO have published recommendations for the prevention and control of COVID-19 for healthcare workers ^11,12^. Knowing, having positive attitudes toward these guidelines, and adhering to the practice of these measures could be essential to avoid preventable deaths and facilitate outbreak management. In our study, we assessed the knowledge, attitudes, and practices of SARS-cov2 infection among Ecuadorian final-year medical students toward the diagnosis, treatment, prevention, and prognosis of COVID-19 in order to identify the knowledge gaps, perceptions, and behavior patterns which could guide the design of better medical education curricula on COVID-19.

## Methods

### Setting and participants

A cross-sectional, online survey was fielded from April 6 to April 20. Participants were students currently enrolled in their final year of medical school in Ecuador. The secretary of the Association of Ecuadorian Faculties of Medical and Health Sciences (AFEME, in Spanish) sent the survey by email to the deans of all 22 universities with a medical school in Ecuador^13^ for distribution to all final-year medical students at each school. The survey was also sent to eligible participants by the coauthors’ Facebook and WhatsApp (V-M.P., R-S.C., B-B.G.).

### Survey development and measures

Data were collected using a 33-item online survey to evaluate medical students’ knowledge, attitudes, and practices (KAP) around COVID-19. A novel survey instrument was developed for this study using items adapted from information about COVID-19 published by the CDC and the WHO ^11,12^ as well as items used in previous COVID-19 surveys ^16,17^. The survey was developed and fielded in Spanish and translated later into English for reporting purposes. The survey was fielded using Google Forms, an online, cloud-based survey administration application. In order to minimize missing data, respondents were required to complete each item in order to proceed to the subsequent items. The full survey instrument is available in the Supplementary Material.

The questionnaire was reviewed for face validity by three experts in medical education, infectious diseases, and biostatistics to identify key issues that may be relevant to final year medical students during the outbreak. After incorporating expert feedback, we pilot-tested the survey instrument online with a group of 30 final-year medical students. During this process, students completed the survey in full and then were interviewed by three members (L-N. E., V-M.P., R-S.C.) of the research team to elicit their feedback and suggestions for improvement. The 30 students who completed the pilot-testing did not participate in the final survey, and the responses collected during pilot-testing were not included in the final analysis.

### Domain: Knowledge

The knowledge domain was composed of 23 questions to evaluate students’ knowledge about COVID-19, including its virology, diagnosis and clinical management, prevention, and relevant infection control measures. For all questions in this domain, respondents were asked to respond ‘true’, ‘false’ or ‘not sure’. A correct answer was assigned 1 point and an incorrect answer or a ‘not sure’ response was assigned 0 points. For each respondent, we categorized a knowledge score of ≥ 16 points out of 23 possible points (≥ 70% correct) as “high knowledge”. Scores of < 16 points (< 70% correct) were categorized as “low knowledge”. The cut-off point is based on the academic approval regulations of the Universidad Central del Ecuador, which considers a demanding cut-off point of 70% of the final grade, which is the minimum necessary to consider a high level of knowledge ^18^.

### Domain: Attitudes

In the attitude domain, students were asked four questions about their opinions toward volunteering at a health facility during the COVID-19 outbreak, and disease control in hospitals in Ecuador. All questions in the attitude domain had “yes” or “no” response options.

### Domain: Practices

Finally, the practices domain was composed of six questions regarding the students’ use of guidelines, trainings and conferences, hand washing, and scientific searching for information (including types of sources consulted). This domain included four questions that required a ‘yes’ or ‘no’ response and two single-select multiple choice questions.

### Demographic characteristics

In addition, we collected demographic information from respondents, including age, sex, name and location of their university, and the current hospital and department where students were being trained.

### Data management

Demographic characteristics data were managed as follows: universities were classified as public and private according to the secretariat of higher education science and technology (Senescyt)^19^ from Ecuador; and hospital were grouped as public and private subsystems. The public system is comprised of facilities run by the Ministry of Public Health, the Ecuadorian Social Security Institute (which includes Rural Social Security, the Armed Forces, and the National Police), and the health services of some municipalities. The private system is comprised of health insurance companies and prepaid plans for medicine providers ^20^. The rotation department was classified into clinic, surgical, gynecology, or pediatrics, and the cities were grouped depending on the sales income reported in billions of dollars in 2018 according to INEC^21^: Quito (more than 50 billion), Guayaquil (between 50 billion and 10 billion), and others (less than 10 billion).

### Statistical methods

Descriptive statistics were computed using number and percentages for all categorical variables and median and interquartile ranges for all numerical values. The normality of each numerical variable was evaluated with the Kolmogorov Smirnov test. Data were analyzed using IBM-SPSS version 19 (IBM, Armonk, NY, USA) and P<0·05 was considered statistically significant.

### Ethical approval

Consent from the participants was gained at the start of the questionnaire with an explanation of the aim of the study. Participants could continue to the full questionnaire only after responding (electronically) to an ‘Agreement of Participation’ form. All questionnaires were anonymized, and no identifiable data was requested.

## Results

After contacting approximately 3000 final year medical student for participation, 309 students completed the survey and were included in the analysis for this study. Descriptive characteristics of the study sample are reported in Table 1. A majority of participants were women (56.3%), between 23 and 25 years old (74.1%), studied in Quito (61.4%) and in a public university (68%). Nearly all (94%) students were currently training in a public hospital.

**Table 1.** Demographic characteristics.

| Total participant (n = 309) |  | n | (%) |
| --- | --- | --- | --- |
| **Age** |  | median: 24, IQ: 16-19 |  |
| **Sex** |  | ... | ... |
|  | Female | 174 | (56.3%) |
|  | Male | 135 | (43.7%) |
| **Study city** |  | ... | ... |
|  | Quito | 190 | (61.4%) |
|  | Guayaquil | 19 | (6.14%) |
|  | Others* | 100 | (32.46%) |
| **University** |  | ... | ... |
|  | Public | 210 | (67.9%) |
|  | Private | 99 | (32.1%) |
| **Hospital type** |  | ... | ... |
|  | Public | 291 | (94.18%) |
|  | Private | 18 | (5.82%) |
| **Department** |  | ... | ... |
|  | Surgery | 38 | (12.3%) |
|  | Pediatrics | 51 | (16.5%) |
|  | Gynecology | 54 | (17.4%) |
|  | Clinic | 166 | (53.8%) |

\* Other cities included the following respondents: Cuenca: 34, Ambato 23; Manta: 18; Esmeraldas: 8; Tulcan: 5; Ibarra: 4; Riobamba: 3; Latacunga: 1; Loja: 1; Machala: 1; Quevedo: 1; Santa Elena: 1.

### Knowledge about COVID-19

Across the entire study sample, the median COVID-19 knowledge score was 17 points out of a possible 23 points (IQR: 16,19, range: 12-22). The vast majority of participants (88%) attained a high knowledge score (≥16). Out of the 309 respondents, 8% scored 20-23 of 23 possible knowledge points, 80.6% scored 16-19 points, and 11.4% scored 12-15 points.

Regarding each subtopic from the set of knowledge questions, a minority of respondents (31.4%) knew the subgenus to which the virus belonged, while 68.6% (212/309) of students were aware of the basic structure of the virus. The vast majority (91.9%, 284/309) of respondents were aware that close contact with infected patients was the primary mode of transmission and that patients with the disease could be asymptomatic, 98.1% realized that cough, fever, fatigue were the typical symptoms of the infection, and 96.8% knew that realtime polymerase chain reaction (RT-PCR) testing of a respiratory specimen was used to diagnose COVID-19. Also, 97.1% of respondents successfully answered that supportive treatment was the only treatment available for patients with COVID-19 and 90.3% knew that hand washing with an alcohol-based hand sanitizer and the use of a mask are the primary measures for preventing transmission. Other questions are detailed in Table 2.

**Table 2.**
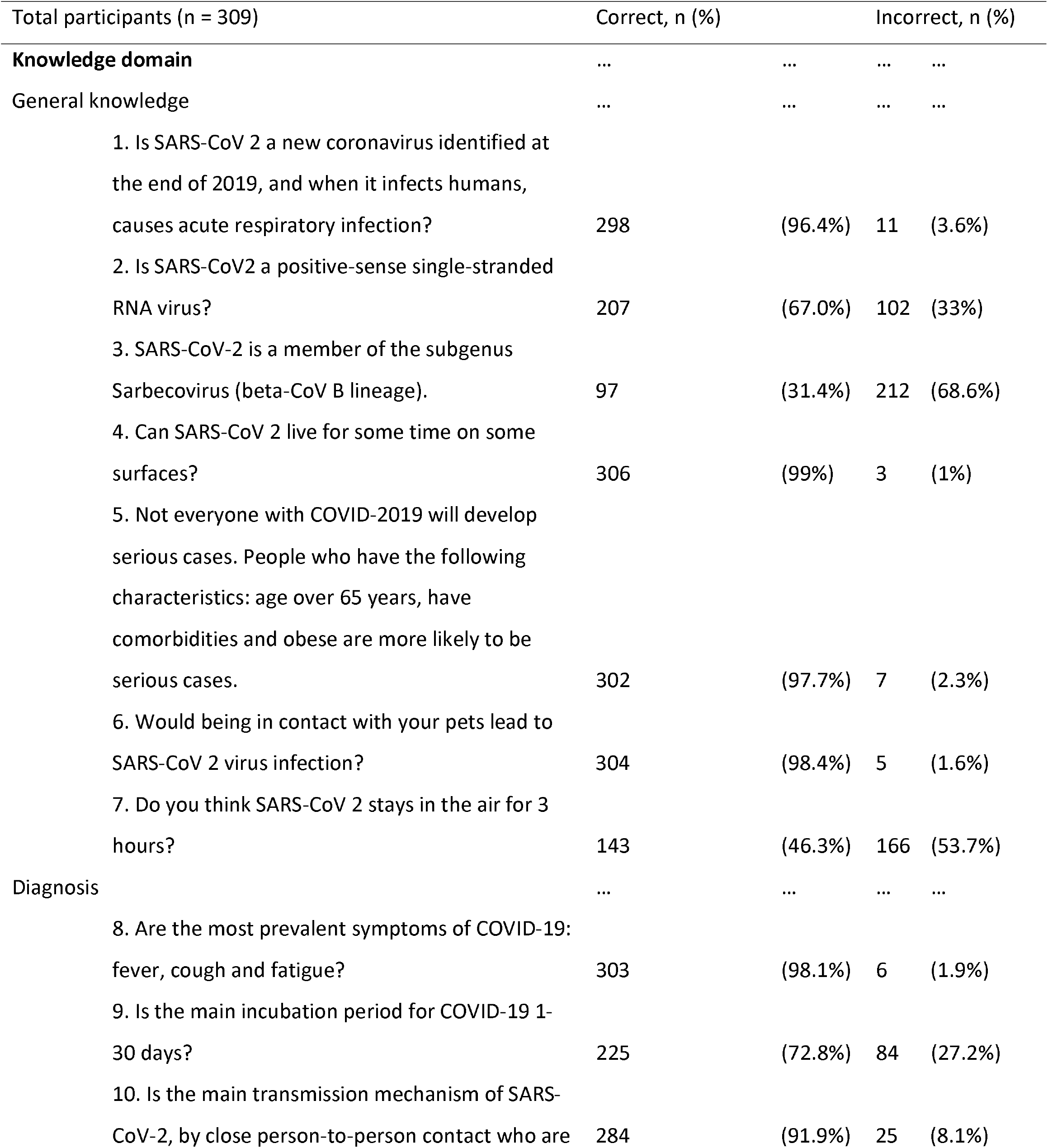
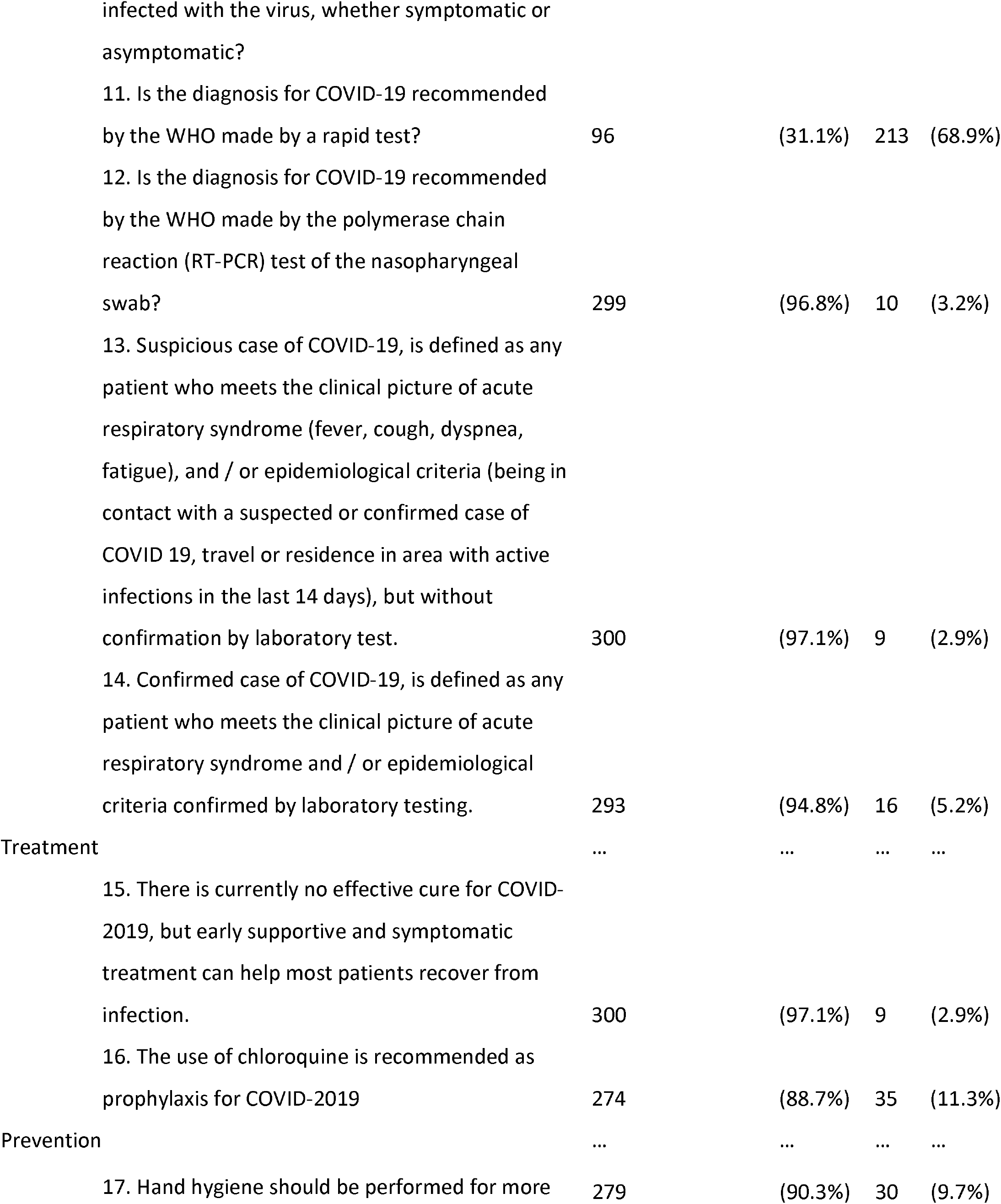
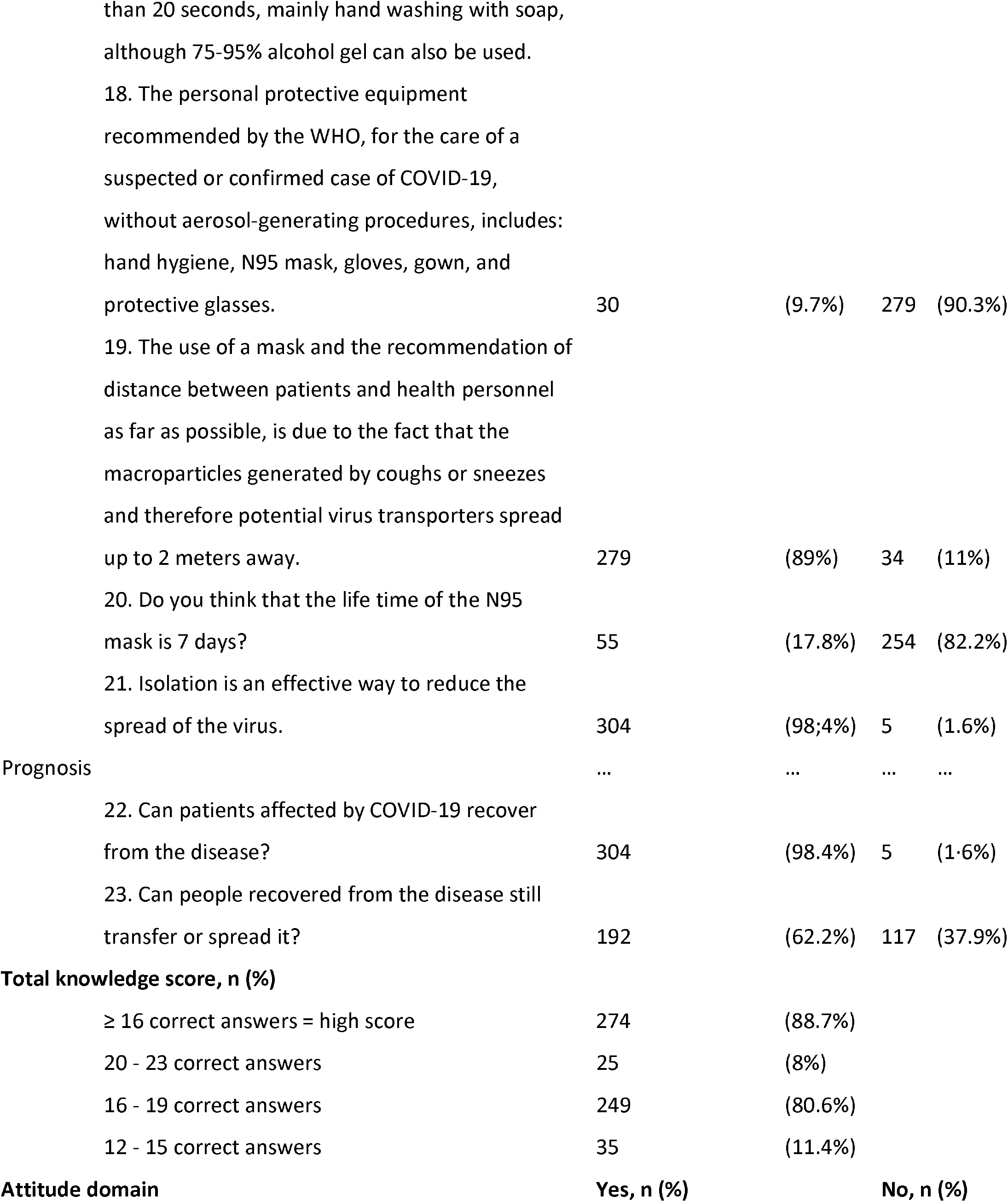
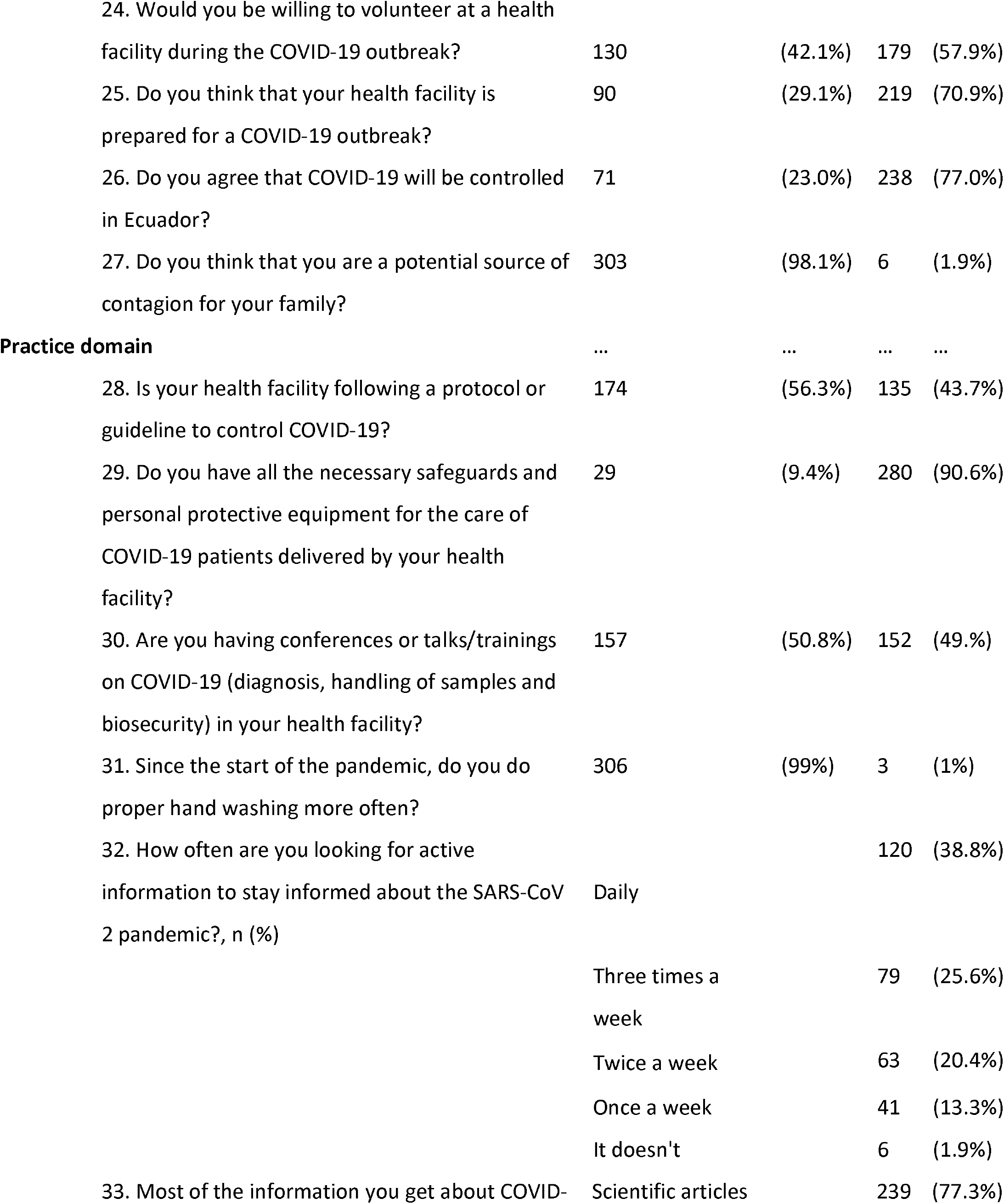
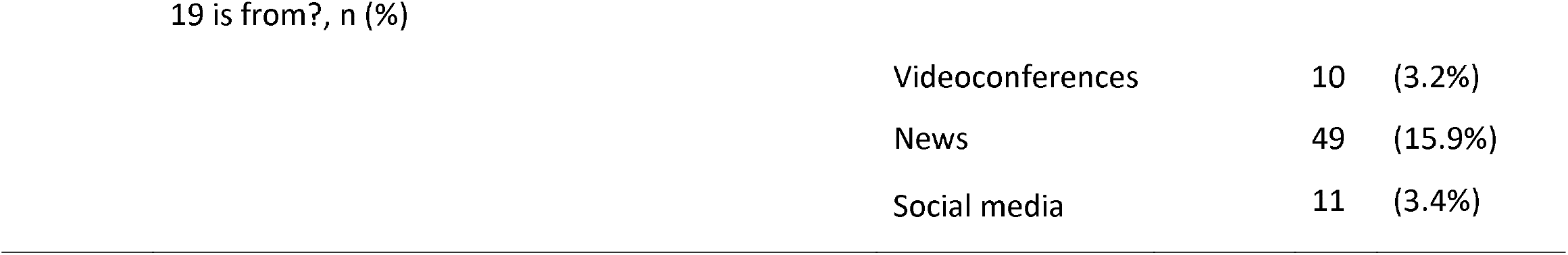
Descriptive statistics for survey responses.

### Attitudes toward COVID-19

More than one half of respondents had a negative attitude toward volunteering at a health facility during the COVID-19 outbreak (57.9%), and a majority of students reported that they did not have confidence that Ecuadorian health facilities and Ecuador in general could win the battle against COVID-19 (70.9% and 77%, respectively). The vast majority of respondents reported believing that they are a potential source of contagion for their families (98.1%).

### Practices related to COVID-19

Only 50.83% of students reported they had any formal talks/training related to infection control, while 56.3% said their health facilities were following a guideline to control COVID-19.

A small number of students reported they received all the necessary safeguards and personal protective equipment (9.4%). Nearly all students said they performed proper hand washing frequently (99.0%). A majority of respondents reported that they obtained information on COVID-19 from reliable sources (77.3% from scientific articles and 3.2% from videoconferences). More than one half of participants reported that they sought updates on the disease three times a week or more (64.4%).

## Discussion

COVID-19 is a global pandemic that involves the entire population, especially health personnel. Final-year medical students are a critical piece of the health care infrastructure, because their role is no longer to merely observe, but to be actively involved in patient care responsibilities ^22–24^. Encouragingly, in our study we found that the majority of final-year medical students had a high level of knowledge about COVID-19 virology, transmission, and management. However, attitudes about the outlook of and personal willingness to volunteer during the COVID outbreak in Ecuador were largely negative. Furthermore, we found evidence that the delivery of adequate personal protective equipment and implementation of trainings were poor, because 90% of the students indicated that they did not receive PPE and only 49% received training. To the best of our knowledge, this is the first study in not only Ecuador but Latin America examining KAP towards diagnosis, treatment, prevention, prognosis, and education of COVID-19 among final-year medical students.

The high levels of knowledge observed in our study is consistent with findings from recent surveys of HCWs in other areas of the world ^16,17,25^. However, it is difficult to directly compare our findings with those of previous studies because of varying definitions for establishing a “high” level of knowledge. This high score could be attributed to the severity of the pandemic and the overwhelming volume of articles, online conferences, and news reports related to this public health emergency. However, a high score in theoretical knowledge is not *sine qua non* of good medical practice, because there are elements related to clinical practice and patient care that such evaluations cannot measure. For that reason, appropriate training is urgently necessary. Unfortunately, this need appears to be unmet by Ecuadorian health facilities and universities, given that almost half of final-year medical students in our study sample reported a lack of formal training about COVID-19. Although we do not have direct evidence of the magnitude of training deficits in other countries, the similarities in medical education models throughout Latin America make it reasonable to assume that lack of COVID-19 training may be prevalent among other countries in the region. This represents a concerning flaw and an important missed opportunity in the global fight against COVID-19. Future studies should assess these deficiencies in more detail and among other Latin American countries in order to identify and improve on such training gaps.

Evidence from Denmark, which added the use of a novel digital platform to its undergraduate medical curriculum as a response to COVID-19, suggests that timely changes to medical education can improve national responses to the pandemic ^26^. Experiences like these demonstrate that Ecuadorian medical students, too, may benefit from additional opportunities to learn about COVID-19 management during this crisis. Findings from our study suggest that Ecuadorian final-year medical students, despite being the future of Ecuador’s front line of health care personnel, will not be adequately prepared to face the challenges of COVID-19 in daily clinical practice. This is particularly important because medical students are part of the solution to cover and complement the shortage of HCWs that exists in Ecuador. According to the latest data reported by INEC^27^ in 2018, there are 23 doctors and 14 nurses per 10 000 inhabitants, equal and less than the minimum established by the WHO ^28^ to provide essential health services to the population (23 skilled HCWs per 10 000). The number of these “new doctors” (3 000) added to the existing doctors in Ecuador (39 908)^11^, would raise the number of doctors per 10,000 inhabitants to 25.2, higher than the minimum established by the WHO. Hence the importance that, depending on the development of a new curriculum, the training of medical students would allow them to cover nursing, telemedicine and social education tasks, which would have a positive impact in the fight against COVID-19, as demonstrated in other countries where, to avoid the collapse of the health systems, the participation of medical students was requested to collaborate in the triage and/or management of suspected or confirmed COVID-19 patients ^29,30^.

Based on similar studies on influenza and other infectious disease in Latin American medical students^31,32^, where students had high level of knowledge of the disease and were pessimistic about possible government actions, the finding of a high COVID-19 knowledge in Ecuadorian final-year medical students and their negative attitudes were expected. Perhaps the lack of adherence ^33,34^ to social distancing measures, the lack of isolation of positive cases (which is difficult in a country where cultural and economic inequalities have led people from informal jobs or underemployment to not follow the recommendations), decades of poor political programming (reflected in the education budget reduction ^35^), and the lack of guidance and clinical supervision have discouraged final year medical students to work as volunteers in a health facility during the COVID-19 outbreak, and led them to believe Ecuador will not control the COVID-19 outbreak. Furthermore, the absence of official and unambiguous statements on indemnity, expected roles and responsibilities, and contractual agreements would make it difficult for students to participate in Ecuador. Additionally, these students’ negative attitudes could be the result of the shortage of protective equipment and medical supplies ^3^, considering we found that 90.6% of students said they do not have appropriate personal protective equipment.

We hope that the results of this study encourage the development of public health policies that improve the decision-making process against a disease without borders and promote and inform the creation of evidence-based national training curriculum against COVID-19, aimed at preparing and educating medical students. Moreover, we believe that having an adequate level of knowledge of COVID-19 will allow this type of program to be implemented easily, which will be of great benefit during this pandemic and for the future.

## Limitations

Our study has several limitations that should be discussed. First, the non-random sampling of this study could be a source of self-selection bias. Although it is not possible to predict the direction of this bias, we believe that it is reasonable to expect that knowledge could be overestimated if those with lower knowledge chose not to participate. Similarly, those that were more optimistic about the situation may have been more inclined to participate, which could have led to underestimated negative attitudes and inadequate behavioral patterns. Future studies could address this limitation by selecting a random sample of medical students; perhaps through the use of institutional e-mails. Social desirability bias is another potential limitation that could have affected attitudes and behavioral pattern responses as a result of the self-report nature of the survey. In other words, it is possible that negative attitudes and inadequate behavioral patterns are underestimated due to respondents having a desire to mark what they consider to be “socially acceptable” responses. However, the use of an anonymous online survey should have mitigated the risk of this bias. Future studies could also avoid this bias by implementing direct observation of practice in order to get more accurate estimates of behavioral patterns. Despite these limitations, we consider this study to be a solid approximation of knowledge, attitudes, and practice of Ecuadorian medical students that can help to inform their training needs and, subsequently, can be used to design a specific curriculum about COVID-19 that could act as an important healthcare resource.

## Conclusions

Like many middle-income countries, Ecuador does not have operational readiness capacities or a definitive actionable plan to combat a pandemic at the scale of COVID-19^36^. The high level of knowledge of final year medical students is an important resource that Ecuador will soon be able to tap, but we must strengthen, improve and advance these students’ training to prepare them for the fight against COVID-19, and to address their concerns that were raised in this study. Now is the time to develop plans for making the transition as safe as possible for students, medical teams, and most importantly, for patients. Creating a national curriculum may be one of the most effective ways for all students to be trained. Students can receive online training specific to COVID-19 and disaster preparedness, and healthcare systems can develop workflows that integrate students where they can be most effective.

## Data Availability

The datasets generated during and/or analysed during the current study are available from the corresponding author on reasonable request.

## Author Approval

All authors have seen and approved the manuscript.

